# Prevalence and associated factors of chronic kidney disease among adults with type 2 diabetes mellitus in Sub-Saharan Africa: A systematic review and meta-analysis

**DOI:** 10.64898/2026.07.07.26357494

**Authors:** Kibrom Aregawi Amare, Girmay Belete Berhe, Gebrehiwet Tesfay Yalew

## Abstract

**Introduction:** Chronic kidney disease (CKD) in adult patients with type 2 diabetus mellitus (T2DM) is a serious public health challenge and continues to be a major source of morbidity and mortality in Sub-Saharan Africa (SSA). The burden of CKD among T2DM patients in SSA has been documented in several studies, but the results remain heterogeneous, and there is limited comprehensive data on its overall prevalence and factors associated with the diseases. Therefore, this study aimed to estimate the pooled prevalence of CKD and assess its associated factors among adult patients with T2DM in SSA.

**Methods:** This systematic review and meta-analysis was conducted and reported according to PRISMA 2020 guidelines, and the study protocol was registered in PROSPERO (CRD420261411707). A comprehensive search of relevant studies was conducted from PubMed, Google Scholar, Scopus, Cochrane Library, and EMBASE published between January 2000 and December 2025 in SSA. Data were analyzed using Stata version 17. A random-effects model was used to estimate the pooled prevalence of CKD among adults with T2DM patients. Heterogeneity was assessed using Cochrane’s Q and I² statistics with visual inspection through forest and Galbraith plots. Publication bias was evaluated using a funnel plot and Egger’s test. Subgroup and sensitivity analyses were also performed.

**Results:** Out of the 3702 study participants from 16 included studies, the estimated pooled prevalence of CKD among adult T2DM patients in SSA was 35.8% (95% CI: 27.1–44.4), indicating significant heterogeneity (I²=97.25%, p<0.001) across the studies. Subgroup analyses on the pooled prevalence of CKD on the basis of different diagnostic criteria were conducted. Modification of diet in renal disease (MDRD) reported a prevalence of 34% (26, 43), the chronic kidney diseases epidemiology collaboration (CKD-EPI) reported a prevalence of 39% (21, 57), and the Cockcroft-Gault (CG) method reported a prevalence of 34% (12, 56). In addition, older age (OR = 2.27, 95% CI: 1.19–4.33) and longer diabetes duration (OR = 1.90, 95% CI: 1.07–3.40) were factors significantly associated with the prevalence of CKD.

**Conclusion:** The prevalence of CKD in SSA was high, affecting nearly one in three adults with T2DM patients. In addition, factors such as older age and longer diabetes duration significantly contributed to the association with CKD. To lessen this issue, focused public health actions are strongly advised, such as screening, education, and awareness campaigns.

**Systematic review registrations:** PROSPERO (2026: CRD420261411707).

## Introduction

Chronic kidney disease is a long-term disease with an abnormal alteration in kidney function or structure, characterized by a lower glomerular filtration rate (GFR), which is less than 60 ml/min/1.73 m², and the presence of proteinuria or both for more than three months. This condition causes the kidneys to slowly lose their ability to filter waste and extra fluid from the blood. It is primarily a microvascular complication of T2DM and is associated with high morbidity, mortality, and cardiovascular problems (1). Diabetes mellitus is currently experiencing a significant increase in both prevalence and incidence worldwide, particularly in low- and middle-income countries, including those in SSA (2).

Patients with T2DM with long-term hyperglycemia, hyperlipidemia, hypertension, diabetic retinopathy, and additional anomalies related to metabolism are more likely to develop CKD (3). Delayed diagnosis, limited access to health institutions, insufficient screening programs, and poor glycemic management may all contribute to the burden of CKD among diabetes patients in SSA. Improving the capacity of health systems to provide service to address the common cardiovascular diseases risk factors, including hypertension and high cholesterol, besides lowering glucose, is key to preventing diabetes-related kidney diseases in low- and middle-income countries (4).

The pathophysiology of diabetes-related CKD is a multidimensional process. Initially, it involves a synergy of hemodynamic and metabolic abnormalities. As the disease progresses, this hemodynamic and metabolic stress triggers structural changes and cellular damage. This damage, in turn, activates chronic inflammatory pathways; ultimately, these processes shift the tissue toward irreversible tubulointerstitial fibrosis, where the kidney’s structural and functional integrity is degraded, leading to a sustained decrease in the GFR and, eventually, renal failure (5).

In 2025, approximately more than 589 million adults were living with diabetes worldwide. Nearly 25 million adults are in the Africa region, largely in SSA (2). Most of the cases are T2DM, accounting for about 90–95% of all diabetes cases (6). Of this, CKD affects approximately 27% of the total T2DM patients globally, with an estimation of 6.5 million in Africa, including SSA (7).

There are several studies on the prevalence and associated factors of CKD among T2DM patients in different countries of SSA. However, the results are inconsistent and vary considerably among the study settings. Moreover, there is limited evidence and data on the pooled prevalence and associated factors of CKD among patients with T2DM in SSA. Therefore, the current systematic review and meta-analysis aims to determine the pooled prevalence of CKD and associated factors among adults with T2DM in SSA.

## Review questions

What is the estimated pooled prevalence of CKD among adults with T2DM in SSA? What factors are associated with the prevalence of CKD among adults with T2DM in SSA?

## Methods

### Protocol and registration

This systematic review and meta-analysis was conducted and reported according to the Preferred Reporting Items for Systematic Reviews and Meta-Analyses (PRISMA 2020) guideline (S1) (8). The study protocol was registered in the international database of the Prospective Register of Systematic Reviews (PROSPERO) with registration number CRD420261411707 prior to commencement of formal screening and data extraction.

### Eligibility criteria

Observational studies conducted among adults aged 18 years and above with T2DM in SSA reporting the prevalence of CKD and its associated factors, published in English, were included. However, review articles, editorials, letters, conference abstracts lacking full text, case reports and case series, studies outside SSA, studies without extractable prevalence data, and studies limited to type 1 diabetes mellitus; and duplicate publications were excluded from the review.

### Study design and search strategy

This study was carried out to determine the prevalence of CKD and its associated factors among adults with T2DM using published articles in SSA. A combination of Medical Subject Headings (MeSH) and free-text terms was used to search in PubMed, Google Scholar, Embase, Cochrane, and Scopus. The relevant published studies were identified from January, 2000, to December, 2025. Example PubMed search strategy: (“chronic kidney disease” OR “CKD” OR “renal impairment” OR “renal insufficiency” AND (“type 2 diabetes mellitus” OR T2DM OR “diabetes mellitus” OR “DM” AND (“prevalence” OR “burden“) AND (“associated factors” OR “predictors” OR “determinants“) AND (“Sub-Saharan Africa” OR “SSA“)). Each electronic database had its own search strategy.

### Variable definition and measurement

The main outcome is the pooled prevalence of CKD among adults with T2DM. CKD was diagnosed according to study-defined criteria, which include the KDIGO guideline: an estimated GFR of less than 60 mL/min/1.73 m² or the presence of albuminuria/proteinuria, or both. Associated factors of CKD among adults with T2DM are the secondary outcome of the study.

### Study selection and quality assessment

All retrieved studies were exported to EndNote v.20 software, and duplicates were removed. The titles and abstracts were screened by two independent reviewers (KA and GT). Reviews of full-text articles deemed potentially eligible were retrieved and assessed for final inclusion. Review full texts for eligibility; disagreements were resolved through discussion or third-reviewer (GB) consultation. The study selection process was presented using a PRISMA flow diagram. The Joanna Briggs Institute (JBI) critical assessment checklist for prevalence and observational studies was used to evaluate the methodological quality of all included research. Studies were categorized as having a low, moderate, or high risk of bias. High-quality studies were included in the study if they received a score above 50% on the quality assessment checklist.

### Data extraction

A Microsoft Excel spreadsheet was used to extract the data. This tool was used to retrieve the following information: author name, publication year, country, study design, sample size, mean age, male and female percentage, number of CKD patients, and technique for defining CKD. The data is extracted independently by two reviewers, KA and GT. A third reviewer (GB) was consulted in case of any discrepancies.

### Statistical analysis

Stata version 17 statistical software was used for data entry and analysis. Pooled prevalence was estimated using a Restricted Maximum Likelihood (REML) effect model. Galbraith plots and I-squared statistics were used to evaluate the heterogeneity, which showed 0% (none), <25% (low), 25% to 75% (moderate), and >75% (high) heterogeneity (9). The associations between variables and the prevalence of CKD were determined using odds ratios. In order to investigate potential sources of variability among the studies, subgroup analyses based on African Union regions, CKD definition techniques, publication year, sample size, and mean age were carried out. Finally, publication bias was assessed using funnel plot and Egger’s test, and sensitivity analyses and meta-regression were carried out to assess the effect of a single study on the overall results by sequentially removing each study to detect possible outliers.

### Ethical approval and consent

Ethical approval and informed consent were not necessary because this study was a systematic review and meta-analysis of data from previously published primary investigations on CKD in SSA.

## Results

### Search results and included studies

A total of 2,565 publications covering research published between January 2000 and December 2025 were found across various electronic databases. Following the EndNote removal of 88 duplicates, 2,477 items were filtered by abstract and title. Sixty-one of these were chosen for full-text analysis. After fulfilling the qualifying criteria, 16 studies were ultimately included in the systematic review and meta-analysis. Throughout the study selection process, the PRISMA 2020 flow diagram was adhered to, ensuring a transparent and systematic identification, screening, and inclusion of relevant studies (Fig 1).

**Fig 1.**
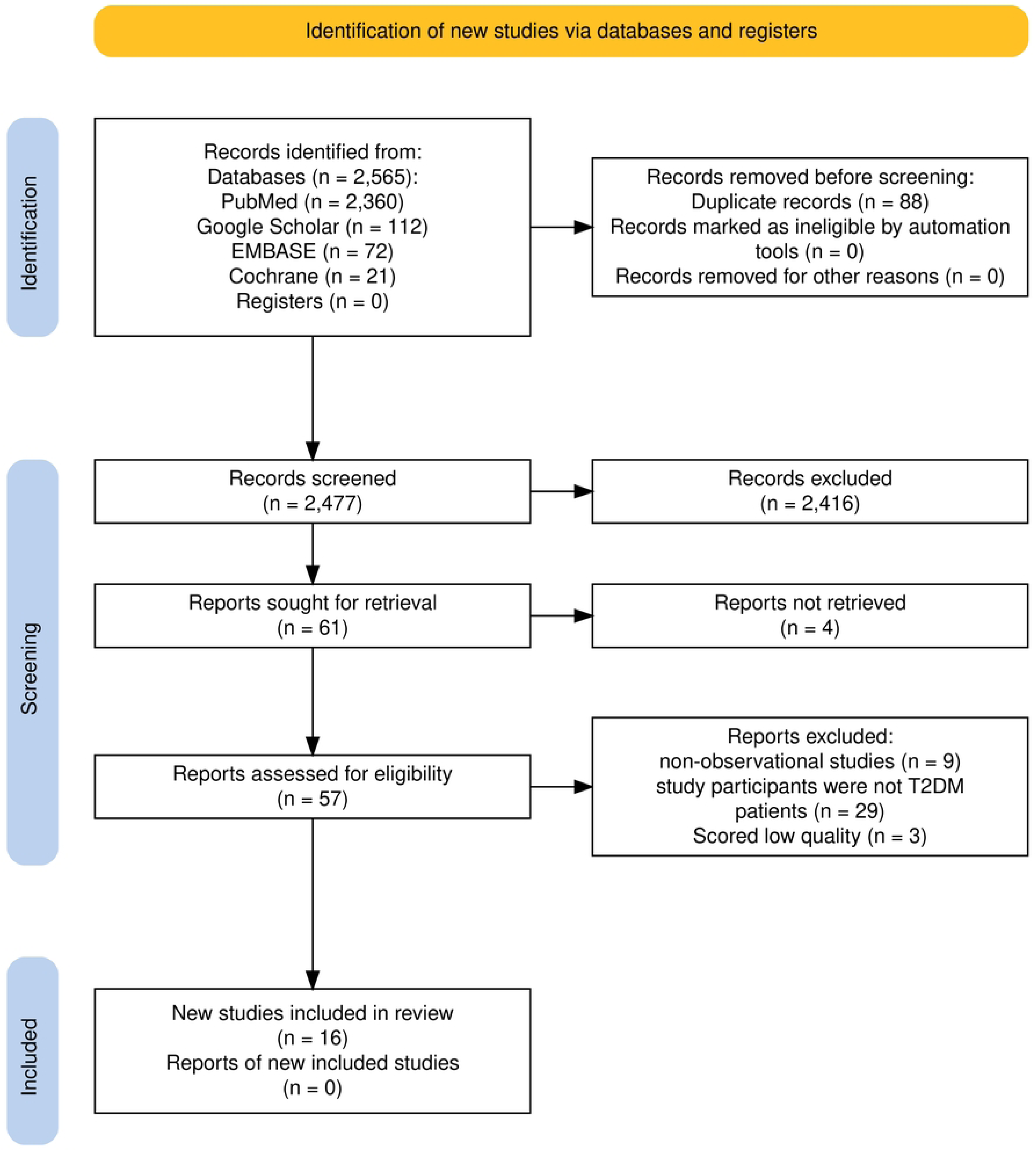
PRISMA 2020 flow chart of study selection process illustrating the identification, screening, eligibility assessment, and inclusion of studies for the systematic review and meta-analysis of the prevalence of CKD and associated factors among adult T2DM patients in SSA. Note. CKD: Chronic Kidney Disease, T2DM: Type 2 Diabetic Mellitus, SSA: Sub-Saharan Africa

### Characteristics of the included studies

In this study, a total of 16 studies with 3702 participants across 8 countries were included and used to estimate the pooled prevalence of CKD among T2DM in SSA. All of the studies were conducted with a cross-sectional study design, and 1502 of the participants were male. The sample sizes ranged from 85 from Cameroon (10) to 408 from Botswana (11). The prevalence of CKD in patients with T2DM was obtained from various countries in SSA: four studies were from Ethiopia (12–15), three from Nigeria (16–18), two each from Kenya (19, 20), Ghana (21, 22), and Cameroon (10, 23), and one each from South Africa (24), Botswana (11), and Uganda (25). These studies reported a total of 1306 CKD cases. Glomerular filtration rate was calculated using the CKD-EPI, MDRD, and CG equations, with six studies using MDRD equations to estimate GFR (14, 18, 20, 23–25). Six articles used the CKD-EPI equation (10, 13, 15, 17, 21, 22); four studies used the CG equation (11, 12, 16, 19). The overall mean age of the study subjects was 56.6 years, ranging from 47 to 63.3. Furthermore, we evaluated the quality of the included studies using JBI critical appraisal checklists. Nearly 62.5% of the studies were categorized as high quality, indicating a low risk of bias and strong reliability of the findings (Table 1).

**Table 1.** characteristics of included studies for chronic kidney diseases among adult type 2 diabetes mellitus patients in Sub Saharan Africa.

| Author & year | Country | Study design | Sample size | Mean Age | Male (%) | Female (%) | Method | CKD cases | Study quality |
| --- | --- | --- | --- | --- | --- | --- | --- | --- | --- |
| Radikara, 2017<sup>(11)</sup> | Botswana | Cross-sectional | 408 | 57.1 | 34.1 | 65.9 | CG | 259 | Low risk |
| Mhundwa et al., 2023<sup>(24)</sup> | South Africa | cross-sectional | 244 | 62.5 | 33.2 | 66.8 | MDRD | 61 | Low risk |
| Otieno et al., 2020<sup>(19)</sup> | Kenya | Cross-sectional | 385 | 63.3 | 34.5 | 65.5 | CG | 150 |  |
| Nibret et al., 2020<sup>(12)</sup> | Ethiopia | Cross-sectional | 162 | 53.9 | 56.2 | 43.8 | CG | 30 | Low risk |
| Mulu et al., 2023<sup>(13)</sup> | Ethiopia | Cross-sectional | 327 | 53 | 43.1 | 56.9 | CKD-EPI | 52 | Low risk |
| Nyamai S., 2014<sup>(20)</sup> | Kenya | Cross-sectional | 200 | 60 | 39.0 | 61 | MDRD | 109 | Low risk |
| Kpene et al., 2024<sup>(21)</sup> | Ghana | Cross-sectional | 141 | 57.4 | 45.4 | 54.6 | CKD-EPI | 99 | Low risk |
| Njonnou et al., 2024<sup>(10)</sup> | Cameroon | Cross-sectional | 85 | 61 | 44.7 | 55.3 | CKD-EPI | 50 | Low risk |
| Georges et al., 2024<sup>(23)</sup> | Cameroon | Cross-sectional | 391 | 59.62 | 32.9 | 67.1 | MDRD | 109 | Low risk |
| Simon et al., 2024<sup>(25)</sup> | Uganda | Cross-sectional | 140 | 53 | 32.1 | 67.9 | MDRD | 47 | Moderate risk |
| Ikem et al., 2023<sup>(16)</sup> | Nigeria | Cross-sectional | 100 | 53 | 56 | 44 | CG | 14 | Moderate risk |
| Tannor et al., 2019<sup>(22)</sup> | Ghana | cross-sectional | 348 | 58 | 43 | 57 | CKD-EPI | 56 | Moderate risk |
| Damtie et al., 2018<sup>(14)</sup> | Ethiopia | Cross-sectional | 119 | 47 | 49.8 | 50.2 | MDRD | 41 | Moderate risk |
| Adem et al., 2024<sup>(15)</sup> | Ethiopia | Cross-sectional | 247 | 49.75 | 61 | 39 | CKD-EPI | 73 | Moderate risk |
| Umar L., 2022<sup>(17)</sup> | Nigeria | Cross-sectional | 261 | 49.6 | 36 | 64 | CKD-EPI | 112 | Moderate risk |
| OKAKA, E., 2015<sup>(18)</sup> | Nigeria | Cross-sectional | 144 | 57.5 | 36.8 | 63.2 | MDRD | 44 | Low risk |

CKD: Chronic Kidney Disease, CKD-EPI: Chronic Kidney Disease Epidemiology
Collaboration, MDRD: Modification of Diet in Renal Disease, CG: Cockcroft-Gault.

### Pooled prevalence of chronic kidney disease in Sub-Saharan Africa

The total pooled prevalence of CKD in SSA was 35.8% (95% CI: 27.1–44.4), according to the random effects model utilizing REML. Every study showed significant heterogeneity (I² = 97.25, p-value < 0.001). The frequency of CKD varied from 14% reported from Nigeria (16) to 70% reported from Ghana (21) among investigations, according to a meta-analysis of 16 research studies involving individuals with type 2 diabetes (Fig 2).

**Fig 2.**
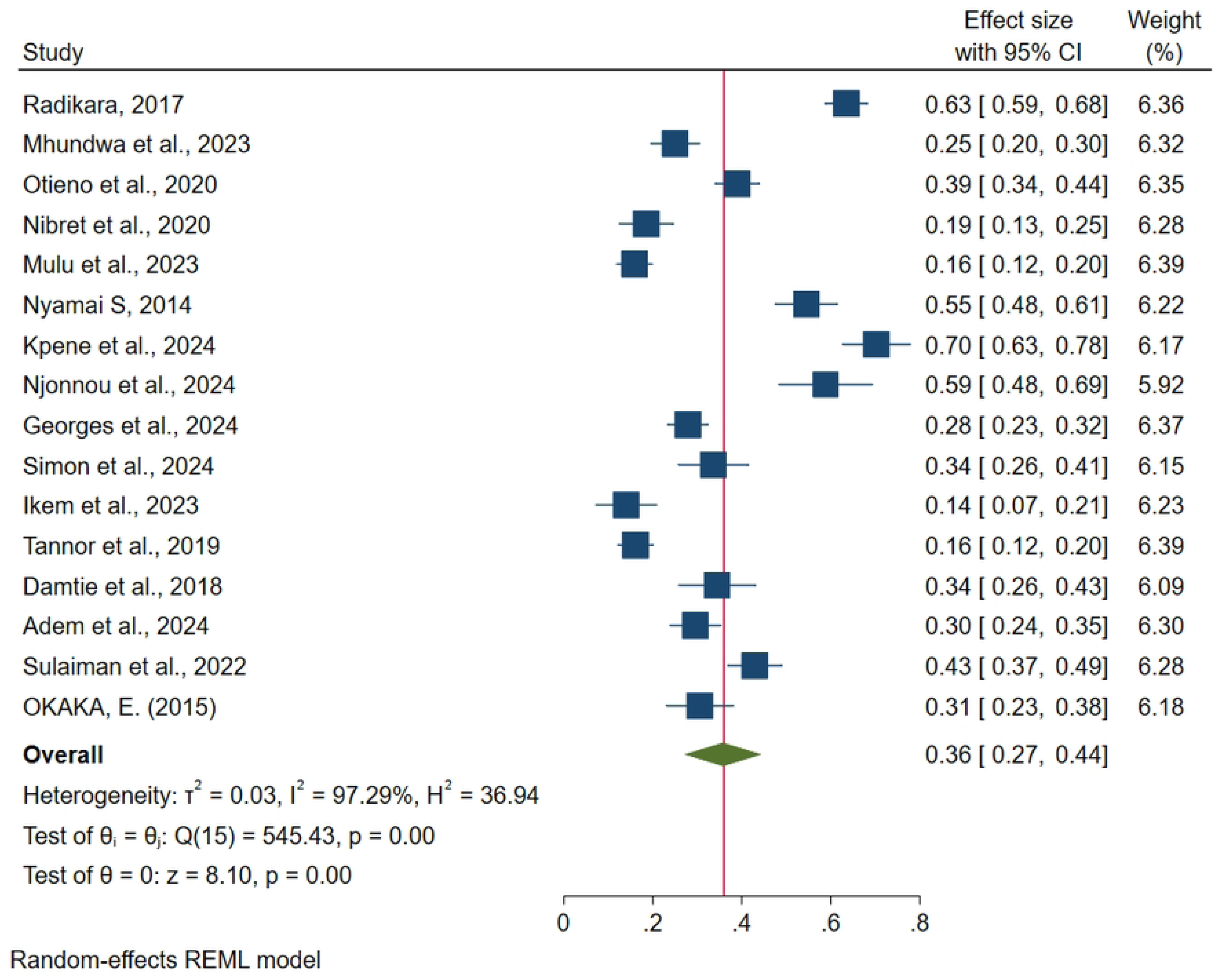
Forest plot of the pooled prevalence of chronic kidney disease among adult type 2 diabetic mellitus patients in Sub-Saharan Africa.

### Publication bias and sensitivity analysis

An Egger’s test and funnel plot were used to visually evaluate the included research for possible publication bias. The funnel plot (Fig 3) appeared slightly asymmetrical, but Egger’s test (β = 8.04, p = 0.087) revealed no evidence of publication bias. A non-parametric trim-and-fill (S1 Table) showed the pooled prevalence remained at 35.8% both before and after adjustment, indicating that publication bias had no significant effect on the total estimate. Additionally, no missing studies were detected. Sensitivity analysis showed stable pooled prevalence not significantly changed by excluding any single study. The result was varied from 33.5% to 37.1%, suggesting the robustness and reliability of the estimate. Additionally, heterogeneity was continuously significant (I²: 96.8%–97.6%), indicating that no single study was responsible for the variability between studies (Fig 4).

**Fig 3.**
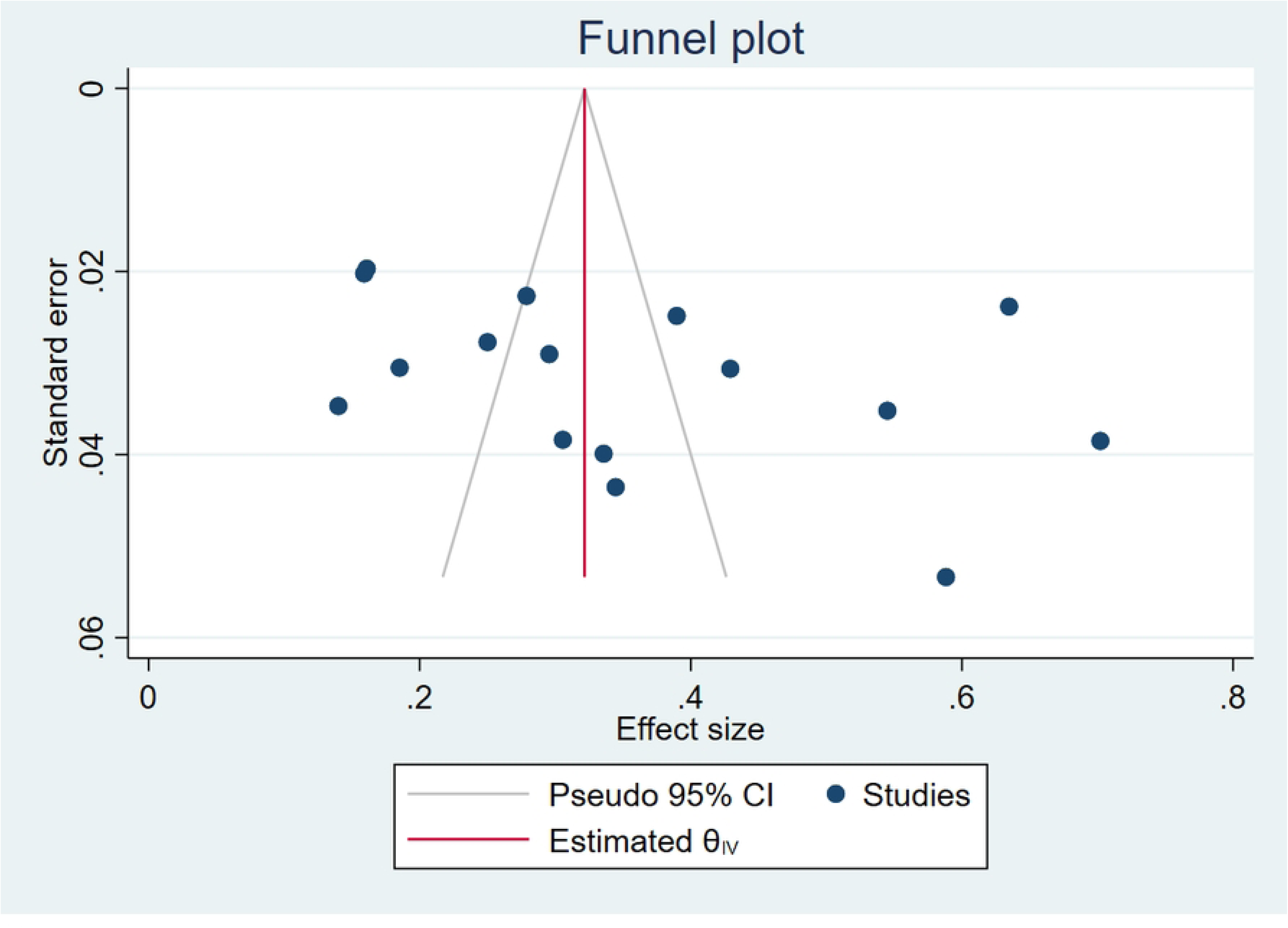
Funnel plot showing publication bias for the prevalence of CKD among adult with T2DM patients. Note. CKD: Chronic Kidney Disease, T2DM: Type 2 Diabetes Mellitus.

**Fig 4.**
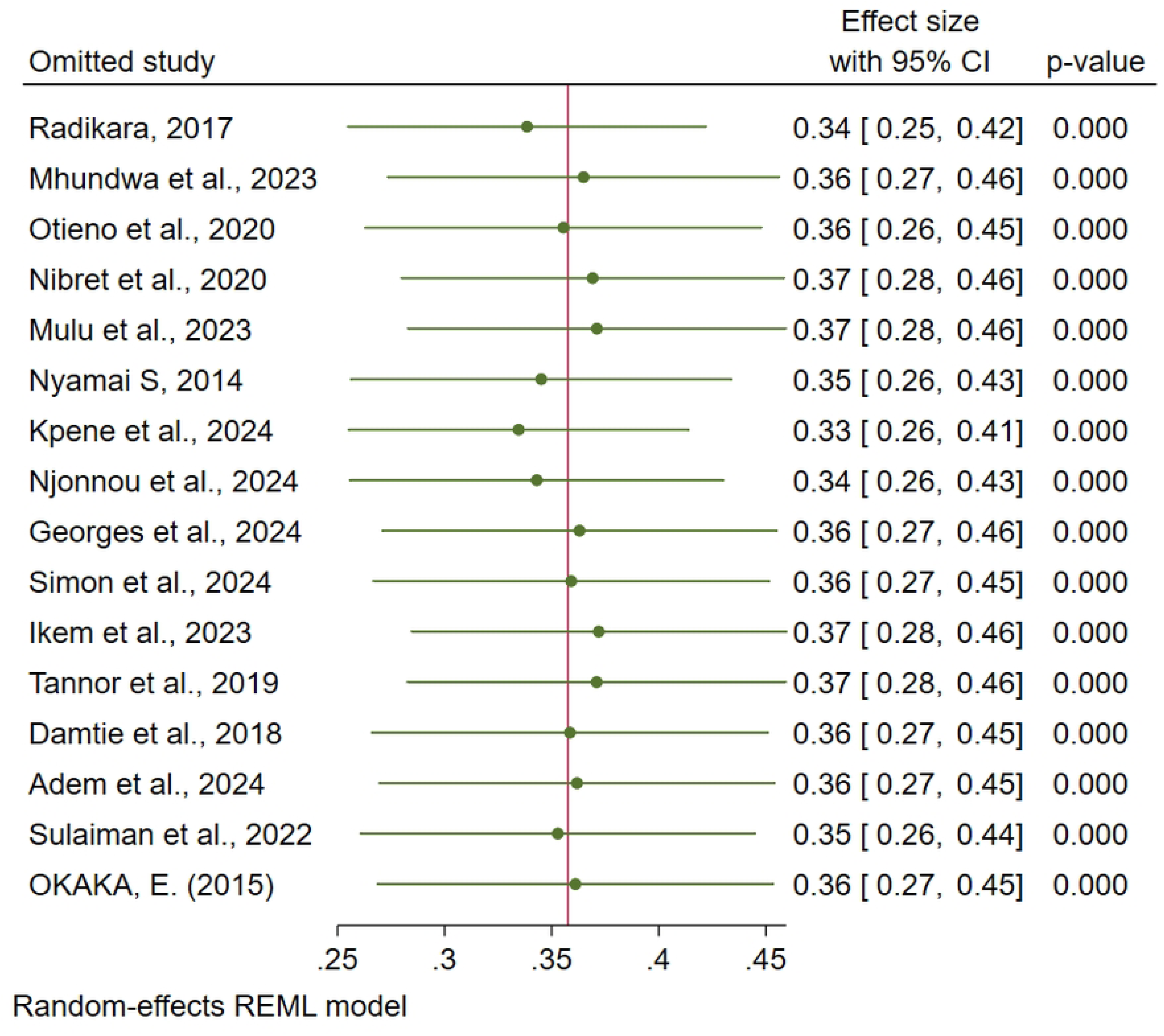
Sensitivity analysis showing the prevalence of CKD among adult with T2DM patients. Note. CKD: Chronic Kidney Disease, T2DM: Type 2 Diabetes Mellitus.

### Subgroup analysis

Statistical tests, including the Cochrane Q test and I² statistics (I² = 97.25%, p < 0.001), along with a forest plot and Galbraith plot (S2), revealed heterogeneity among the studies. Subgroup analyses were conducted based on African Union regions, mean age, publication year, methods used to define CKD, and sample size.

Subgroup analysis by African region was conducted, and the result revealed that Southern Africa and Central Africa showed the highest percentage of prevalence of CKD at 44.3% (95% CI: 6.6–82.0) and 43.0% (95% CI: 12.7–73.3). This is followed by West Africa at 34.7% (95% CI: 14.5–54.8), and the lowest in East Africa at 32.1% (95% CI: 22.4–41.8). The I^2^ ranged from 94.8% to 99.1%, indicating significant variability in all subgroups. However, there was no statistically significant difference in the prevalence of CKD amongst sub-Saharan African countries, according to the test for subgroup differences (Q = 0.78, p = 0.855) (S1 Fig).

Subgroup analysis based on study sample size revealed variations in CKD prevalence. A higher CKD pooled prevalence of 39.2% (95% CI: 25.4, 53.0) and 32.4% (95% CI: 21.5, 43.4) was reported in studies with a sample size of less than 222 participants and studies with 222 or more participants, respectively. However, the difference between the two subgroups was not statistically significant (Qb = 0.57, p = 0.452) (S2 Fig).

The mean age-based subgroup analysis showed differences in the prevalence of CKD. Pooled CKD prevalence was 31.6% (95% CI: 20.1, 43.1; I² = 96.82%, p < 0.001) in studies with a mean age of less than 57 years, and 40.0% (95% CI: 26.9, 53.1; I² = 97.61%, p < 0.001) in those with a mean age of 57 years or older. Though, the two subgroups’ differences were not statistically significant (Qb = 0.90, p = 0.343) (S3 Fig).

The prevalence of chronic kidney disease (CKD) varied according to the subgroup analysis based on the method used for its definition. The CKD-EPI equation showed a combined prevalence of 38.7% (95% CI: 20.7, 56.6; I² = 98.44%, p < 0.001), while the Cockcroft–Gault (CG) equation indicated a combined prevalence of 33.8% (95% CI: 11.7, 56.0; I² = 98.48%, p < 0.001). Studies that employed the MDRD equation yielded a combined prevalence of 34.2% (95% CI: 25.7, 42.7; I² = 90.31%, p < 0.001). However, there was no statistically significant difference observed among the subgroups (Qb = 0.20, p = 0.904) (S4 Fig).

### Meta-regression

The cause of the heterogeneity in the included studies was determined using meta-regression analysis. Effect size was not statistically significantly predicted by the factors of publication year, mean age, or sample size (p > 0.05). None of the factors accounted for the observed between-study heterogeneity (R^2^ = 0%). The I^2^ = 96.83% indicates that the residual heterogeneity still remains considerable (Table 2).

**Table 2.**
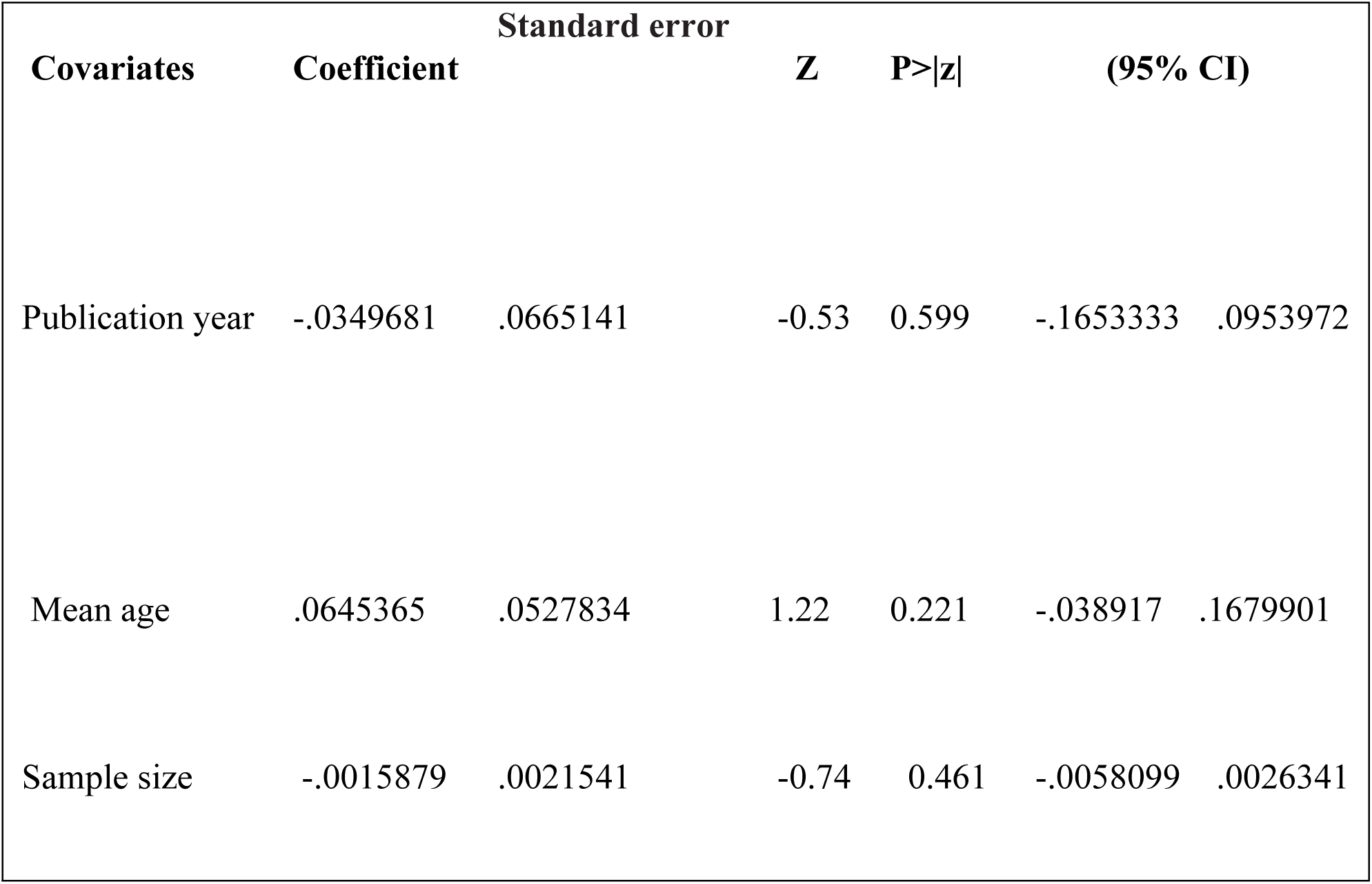
meta-regression analysis results for the pooled prevalence of chronic kidney disease among adult type 2 diabetes mellitus patients in Sub-Saharan Africa.

| Covariates | Coefficient | Standard error | Z | P>\|z\| | (95% CI) |  |
| --- | --- | --- | --- | --- | --- | --- |
| Publication year | -.0349681 | .0665141 | -0.53 | 0.599 | -.1653333 | .0953972 |
| Mean age | .0645365 | .0527834 | 1.22 | 0.221 | -.038917 | .1679901 |
| Sample size | -.0015879 | .0021541 | -0.74 | 0.461 | -.0058099 | .0026341 |

### Associated factors of chronic kidney disease among adult T2DM patients

A random-effects model using REML was applied to estimate the pooled odds ratios for each factors extracted from the included study. The method revealed that older age, longer diabetes duration, and obesity were factors that were strongly associated with CKD. A greater than two-fold increase in the likelihood of CKD was linked to older age (OR = 2.27, 95% CI: 1.19–4.33). In a similar vein, the odds of CKD increased 1.90 times with longer diabetes duration (OR = 1.90, 95% CI: 1.07–3.40). On the other hand, obesity was associated with a protective effect against CKD (OR = 0.29, 95% CI: 0.18–0.47). Nevertheless, poor glycemic management (OR = 1.03, 95% CI: 0.32–3.25) and hypertension (OR = 1.29, 95% CI: 0.30–5.45) did not significantly correlate with CKD.

## Discussions

This systematic review and meta-analysis aimed to determine the pooled prevalence of CKD in SSA. The primary data was collected from 16 different articles that included 3,702 adult with T2DM patients. This study highlighting the substantial impact of renal disorders in this population, and revealed an overall prevalence of CKD was 35. 8% (95% CI: 27. 1–44. 4). This result aligns with findings from a systematic review and meta-analysis conducted in Ethiopia 35.52% (26), and Middle East Region 28.96% (95% CI: 19.80–38.11 (27), as well as with other SSA hospital-based studies like Kenya 39.0% (95%CI, 34.3–44.2) (19), and Uganda 33.6% (95% CI: 26.2-41.9) (25). However, it is higher than the similar study setting in SSA 13.9% (28), worldwide prevalence (27%) (7), Bangladesh 21.3% (29), in northern Thailand 24.4% (21.9–27.0) (30), Spain 27.9% (CI 95% = 25.2 - 30.5) (31), south Africa 25.0% 20.0–30.8% (24). This may be explained by variations in the study population’s characteristics and CKD diagnosis criteria, moreover, the study’s hospital-based design might have made it more likely that patients with more severe illnesses and consequences would be enrolled. The disparity could also be explained by variations in the load of comorbidities, especially hypertension and longer-term diabetes. Furthermore, this study result is lower than the study conducted in Singapore 53% 50.7%−55.3% (32), Tanzania 83.7% 80.0–87.5 (33), Australia 47.1% (95% CI: 45.8–48.4) (34), and Botswana 63.5% (95% CI: 58.7% to 68%) (11). The lower prevalence observed in this study may be due to the method CKD-EPI recent equation to define CKD, which is more precise than the MDRD and CG equation utilized in those comparative studies. used The CKD-EPI, equation used for eGFR calculation was utilized by the majority of the articles included in this study that can lead to yield higher eGFR. This high eGFR potentially resulting in fewer individuals being classified as having CKD. Studies performed at different times might be one reason why prevalence rates that have been declared vary. These studies could show improvements or reductions in how the condition is managed and treated. The meta-analysis found significant heterogeneity between studies and this heterogeneity continued in subgroup analyses and meta-regression. Subgroup analyses based on study region, sample size, mean age, and CKD definition criteria were carried out; however, none of these variables were found to provide a significant explanation for the observed variability. Further confirmation that publication year, mean age, and sample size were not significant predictors of effect size (R² = 0%) came from meta-regression analysis. This implies that diagnostic criteria, genetic predisposition, diabetes treatment regulations, and difference in healthcare service and system are just a few of the unmeasured variables that may be responsible for the heterogeneity. Subgroup analysis based on African region, revealed that Central and Southern Africa have highest frequency. Whereas, the lowest in East Africa. Despite study setting difference, the variation across the region was not statistically significant, suggesting that the burden of CKD among T2DM patients is generally consistent throughout SSA. Similarly, CKD prevalence was higher in studies with older individuals and smaller sample size, while these variations were likewise not statistically significant. Changes in CKD diagnosis methods had no appreciable impact on pooled assessments, demonstrating consistency in how CKD is defined across eGFR equations. Publication bias was checked by egger’s test and trim-and-fill analysis The result revealed no significant publication bias, despite the funnel plot’s had minor asymmetry. The stability of the pooled prevalence after sensitivity analysis gives additional evidence for the results’ robustness since the aggregate estimate stayed essentially the same even when individual studies were omitted. The meta-analysis identified older age and longer duration of diabetes as a significant factors associated with CKD among T2DM patients. The likelihood of developing CKD was more than two times higher in older age adults (OR = 2. 27), most likely due to the age-related decrease in renal function and prolonged exposure to diabetic issues. This finding is supported by research carried out in Ethiopia, which indicated that elderly people with diabetes had an increased risk of developing CKD (35). Additionally, a study in Jordan demonstrated that age is a significant factor influencing CKD prevalence among diabetic patients (36). The KDIGO 2012 Guideline for Evaluating and Managing CKD indicates that the incidence of CKD increases with age because of the natural deterioration of kidney function and the accumulated impact of risk factors like diabetes and hypertension (37). Patients with longer duration of diabetes were 1.90 times more likely to develop CKD than short duration on diabetes, which may reflect cumulative hyperglycemic destruct to micro-vascular structure kidney. This result also supported by a study conducted in Ethiopia, revealed that the risk of CKD significantly increases with the duration of diabetes (38). This is due to prolonged exposure to high glucose level in the blood, which progressively deteriorates the micro-vascular structure through oxidative stress, advanced glycation end-product accumulation, and chronic inflammation. Over time, these processes proceed to glomerular basement membrane thickening, mesangial expansion, and nephron loss, ultimately impairing renal function and increasing the risk of developing CKD (39). Obesity, another factor, showed a protective statistical significance with CKD (OR = 0.29), which could indicate possible survival bias, variations in body weight measurement, or confounding effects in particular investigations. However, the pooled analysis showed no significant relationship between CKD and hypertension or poor glycemic control. The disparities in glycemic evaluation techniques across studies, therapy effects between hypertensive individuals, or changes in measurement may contribute for this. The urgent necessity for early screening and coordinated diabetes-kidney care initiatives should emphasizes due to the high burden of CKD in adult T2DM patients in SSA. Focus should be placed on routinely monitoring renal function, especially in long-standing diabetic patients and older patients. Early diagnostic tools like eGFR estimation and the strengthening of primary healthcare systems have the potential to greatly lower CKD-related morbidity. This study has several strengths, including comprehensive evidence synthesis, the inclusion of multiple SSA countries, the use of rigorous statistical techniques, such as random-effects models by using REML, comprehensive subgroup and meta-regression analyses, forest plots, and sensitivity analysis were employed to improve the validity and reproducibility of the results. However, some limitations include limited geographic representation, use of secondary data, significant heterogeneity that could not be fully explained, due to differences population characteristics, sample size, and reliance on cross-sectional studies, and variability in CKD diagnostic methods.

## Conclusion

This study demonstrates a high burden of CKD among patients with T2DM in SSA, affecting nearly one in three adults with T2DM patients with significant associations observed for older age and longer duration of diabetes. These findings underscore the need for early detection strategies and improved chronic disease management to reduce the burden of CKD in this population.

## Data Availability

No new datasets were generated during the current study. The data supporting the findings of this study were extracted from previously published articles, all of which are cited within the manuscript and its supplementary information files. The extracted data used for the meta-analysis are included within the manuscript and its Supporting Information files.

## Author contributions

Conceptualization: Kibrom Aregawi Amare, Girmay Belete Berhe, Gebrehiwet Tesfay Yalew

Data curation: Kibrom Aregawi Amare, Girmay Belete Berhe, Gebrehiwet Tesfay Yalew

Data analysis: Kibrom Aregawi Amare

Investigation: Kibrom Aregawi Amare, Girmay Belete Berhe

Methodology: Kibrom Aregawi Amare, Gebrehiwet Tesfay Yalew

Project administration: Kibrom Aregawi Amare

Software: Kibrom Aregawi Amare

Writing – original draft: Kibrom Aregawi Amare

Writing – review & editing: Kibrom Aregawi Amare, Girmay Belete Berhe, Gebrehiwet Tesfay Yalew

## Acknowledgements

We express our gratitude to all of the authors of the studies that were part of this meta-analysis and systematic review.

## Supporting information

**S1 Fig. Subgroup analysis of chronic kidney disease prevalence among adult type 2 diabetes mellitus patients, based on African region in Sub-Saharan Africa.**

**S2 Fig. Subgroup analysis of chronic kidney disease prevalence among adult type 2 diabetes mellitus patients, based on study sample size in Sub-Saharan Africa.**

**S3 Fig. Subgroup analysis of chronic kidney disease prevalence among adult type 2 diabetes mellitus patients, based on mean age in Sub-Saharan Africa.**

**S4 Fig. Subgroup analysis of chronic kidney disease prevalence among adult type 2 diabetes mellitus patients, based on the method of CKD definition in Sub-Saharan Africa.**

**S1 Table. Nonparametric trim-and-fill analysis of publication bias**

**S1 Checklist. PRISMA 2020 checklist for reporting systematic reviews and meta-analyses.**

